# Overview of The Current Progress of Facultative Anaerobic Bacteria in Cancer Biotherapy with TNF-α as Main Mechanism of Action

**DOI:** 10.1101/2021.04.03.21254792

**Authors:** Martina Johansson, Fredrik H Nystrom

## Abstract

This review article focuses on the use of infectious bacteria as delivery tool for tumour necrosis factor α (TNF-α), a well-studied cytokine, in the context of immunotherapy for cancer treatment. The tumour targeting properties of certain bacteria strains has been known for decades as well as the tumour catabolizing effect of TNF-α. The combination of these two have been studied in murine models for various types of cancer with promising results. Research in this fascinating field is unfortunately uncommon, thus the number of high-quality articles is limited. Search was done via Google Scholar in combination with PubMed, to increase the coverage and find peer-reviewed, original, and primary research articles.

Key findings show that attenuated or genetically modified species of bacteria have fewer side effects and can be effective in delivering cytokines to tumour sites. TNF-α is produced by macrophages/monocytes during acute inflammation or infection, thus can be triggered by infectious bacteria which in turn induce apoptosis. The cytotoxic effect of the bacteria can be enhanced with localized irradiation. Promising results have been shown in bladder, breast, colon, glial, lung, ovarian, pancreatic, prostate, and renal cancer cells.

The need for better, safer, and more effective cancer treatment is apparent as traditional chemotherapy and radiation can cause a lot of harm for the patient, and not necessarily prolong the lifespan. The success-rate for these treatments vary greatly depending on the type of cancer, but for tumours that cannot be surgically removed the outcome is generally quite poor. A drawback of chemotherapy is that tumours can grow resistant to the treatment while healthy cells continue to be exposed, increasing the risk of severe side effects. Different types of biological therapies are a modern and possibly safer approach, even though immunotherapy comes with substantial risks. Using the innate immune system to fight tumour cells is not always safe, because uncontrolled and excessive release of pro-inflammatory signalling molecules can result in multisystem organ failure and death. This phenomenon is called cytokine release syndrome (cytokine storm) and is one of the major risks of immunotherapy. However, tailored biological therapies have proven their effectiveness for a wide range of cancer types, and the next step in this evolution is to genetically engineer both delivery systems and mechanisms of action. This approach can be combined with the traditional radiation and chemotherapy for increased effectiveness, even if biological therapies as a stand-alone treatment, can be a goal for the future.

## Background

Tumour necrosis factor α (TNF-α) is playing an important role in apoptosis, inflammation, and immunity. Even though its name suggests antitumor properties, it has been implicated in a wide spectrum of other diseases. TNF-α is a member of the TNF superfamily and was the first cytokine to be evaluated for cancer biotherapy.[1] Unfortunately, its clinical use is limited by its toxicity. Currently, TNF-α is administered only through specific drug delivery systems to reduce the systemic toxicity.[2] Several novel techniques focus on engineered bacterial capsules to deliver TNF-α to tumour sites, and this review summarizes these recombinant microbial treatments currently under investigation. Anaerobic bacteria like *Streptococci, Bifidobacterium, E*.*coli, Listeria, Clostridium* or *Salmonella* are some of the bacteria that can escape the bloodstream and reach the chaotic vasculature of the tumour. This gives rise to inflammation caused by the sudden influx of TNF-α in the tumour vessels.[3] In contrast to the much more clinically used TNF-α antagonists, the desired mechanism here is TNF-α agonism as the inflammatory and toxic properties of this cytokine is used to attack and kill cancerous cells.[4]

Many anaerobic bacteria possess inherent tumour-targeting and tumour-killing qualities as well as overcome penetration limitations because of their small size. Anaerobic bacteria proliferate only in the absence of oxygen, necrotic areas, characteristic of solid tumours.[5] *Salmonella* is one of the most effective bacteria for bacteriotherapy, but can grow under both anaerobic and aerobic conditions, thus can colonize healthy tissue as well. Bacteria with such growth adaptations must be attenuated before use. This can be done by genetic modification or by using heat killing.[6]

Using bacteria in cancer treatment is not a new idea, in fact it stems back to 2600BC when Egyptian physicians tried to facilitate the development of an infection at tumour sites to cause regression of the tumour. The concept of bacteria or any biological mechanism was not known at this time, but the idea of letting the body take care of the tumour with the help of natural processes eventually became standard treatment. However, it was not until the late 19^th^ century that the method really caught a reputation. The American oncologist William Coley developed a mixture of heat-killed bacteria that he named “Coley’s toxin” and injected intravenously into his cancer patients.[7] Up until that time infections had been facilitated with primitive methods such as the use of poultice or just by leaving surgical wounds open and contaminated. Coley’s toxin was safer as he used heat-killed bacteria to reduce the danger. The bacteria used was *Streptococcus pyogenes* and *Serratia marcescens*, two strains that are being studied to this day, as tools for novel cancer therapies.

Even though Coley believed that his concoction was highly effective, the scientific evidence was not of today’s standard.[8] Injecting patients with bacteria to induce a systemic infection followed by high fever is potentially dangerous, but so is chemotherapy and other cancer drugs, thus Coley’s toxin was used against many different types of cancer from 1893 to 1963. Because of the mixed clinical results, FDA made it illegal outside of clinical trials after 1963 and it was later entirely replaced by chemotherapy.

There are several rationales proposed for how Coley’s toxins affect the patients, and several of these are being used in bacterial immunotherapy today. TNF-α is one of the cytokines that make up the acute phase reactions involved in systemic inflammation which results in fever and activated macrophages - a type of white blood cells that engulfs and digests anything that does not have the type of proteins specific to healthy body cells on its surface.[9] The problem with cancer cells is that they are generally not recognized as a threat, but rather as the normal body tissue from which they stem, thus being left to proliferate uncontrollably. When macrophages are in defence mode, they are more prone to attack these types of cells, sometimes causing a complete remission of the tumour.[10] This is the goal of immunotherapy in general and bacteriotherapy is a means to achieve that.

Experiments have been performed in vitro and replicated in vivo. The general principle of Coley’s toxins is correct because some cancers are sensitive to the enhancement of the patient’s immune system. The innate immune system manages growth retardation, and the adaptive immune system has the capacity to eliminate the tumor completely. In an exceedingly rare number of cases this has been observed to happen spontaneously.

The problem is that bacteriotherapy is difficult to manage in a clinical setting. Even though it has great potential to yield promising results, it is unpredictable and needs to be strictly tailored to the individual patient. In recent years only one bacteria-mediated tumour therapy (BMTT) is FDA approved and it is Bacillus Calmette-Guerin (BCG), an attenuated strain of *Mycobacterium bovis*, used for the treatment of non-muscle invasive bladder cancer (NMIBC). This bacterium is recombinant expressing a bacterial toxin and induces TNF-α and Il-10 in the bladder, which is proven more effective than chemotherapy thus is currently the preferred treatment for this type of cancer.[11] Although clinical application of cancer bacteriotherapy is far from routine, the approach has a promising future with more genetic engineering and tailor-made non-hazardous bacteria that can treat cancer.

Bacteria’s have many mechanisms of action when it comes to anti-tumour effect for example enhancing or activating the immune system, be useful as carriers for cancer therapeutic agents, release toxic substances, form biofilms, and invade and colonize solid tumours. Most promising mechanism of action seem to be the activation of an inflammatory pathway with significant increase of inflammatory cytokines. It results in tumour growth suppression by the most important mechanism of the immune system against tumour tissue; TNF-α, which has the potential to destroy the vascular endothelial cells and forms a large haemorrhaging area within the tumour.[12]

## Introduction

Bacteriotherapy is an approach that has shown positive effects of tumour regression and inhibition of metastasis. Bacteria can be genetically engineered to alter their ability to release specific compounds or tailor their metabolic pathways. Anaerobic bacteria especially target hypoxic areas of tumours and actively penetrate tumour tissue.[13] Genetically modified bacteria may also be able to lower pathogenicity to the host and increase the antitumor efficacy. In recent years, numerous experiments have had the aim to use genetic modification, to get bacteria to express reporter genes, cytotoxic protein and/or anticancer agents, and tumour□specific antigens. It is known that genetically modified bacteria generally have more significant multiplication in tumours than in normal tissues. [14] Necrotic regions exist only within tumours and not in normal tissue and that’s where anaerobic bacteria thrive.[15] *Bifidobacterium longum* is an example of bacteria that grow specifically within tumours but not within normal tissues.[16] However, staining of tumour tissue reveal that most bacteria cluster together, rather than distribute throughout the necrotic region. The use of *Salmonella typhimurium* and *Clostridium butyricum* have had higher success rate in targeting the entire region, without severe immune responses or toxic side effects. *Clostridium* in general seem to have many possibilities for biological engineering. It can selectively target and lyse tumor cells and intravenous administration of nonpathogenic bacterial strains or spores from nonpathogenic strains of *Clostridium* bacteria can serve as a selective tumor killer of areas overlooked by conventional cancer therapies. In particular, *Clostridium novyi* is a highly mobile spore-forming organism that is extremely sensitive to oxygen and has never been shown to germinate in areas of tumors outside of the hypoxic regions. The bacteria are exquisitely sensitive to oxygen but its spores are stable to oxygen as well as to harsh conditions. Due to its highly motile nature, the bacteria can easily disperse itself throughout the tumor.[5] Studies also found that bacteria can produce antibodies that bind to hypoxia inducible factor 1α (HIF1-α). HIFs are master regulators of cancer cell’s response in an oxygen-deficient environment to sustain their survival and growth.

HIFs activate the transcription of genes involved in cell proliferation, angiogenesis, metabolism, and invasion. These studies are specifically done with Salmonella in murine models. [17, 18] Cytokines released by the bacteria often belong to the TNF-α superfamily and most commonly involve TNF◻related apoptosis◻inducing ligand (TRAL□l), FAS ligand (FAS◻l), and TNFA. These proteins selectively cause programmed cell death via death receptor pathways, activating the apoptotic mediators. [19] There is a link between tumour growth and inflammation as many of the same processes are involved, such as HIFs and cytokines. Macrophages and dendritic cells are the main producers of TNF-α which are markedly increased in tumours targeted by bacteriotherapy and associated with tumour regression.[20] The aim of this review is to examine the current investigative status of TNF-α and bacteriotherapy, as a potential future treatment option for various types of cancer. Especially for treating solid tumours with hypoxic tissue more prone to be resistant to chemotherapy, as it generally requires rich vascularity to be effective.

## Methods

Research on bacteriotherapy is widely popular but bacterial delivery vectors with TNF-α as the main mechanism of action is less common, and that is why Google Scholar was used as a complement to PubMed to get a wider search range. Google Scholar indexes the full text of most peer-reviewed online academic journals and books, conference papers, theses, dissertations, abstracts, patents, and technical reports.[21] The search was focused on peer-reviewed articles written in English, published during a 20-year period from 1999 to 2019. The search words were “bacteria mediated tumour therapy”, “TNF-α” “cancer immune therapy” “oncolytic bacterial therapy” and “microbial oncology therapeutics”. The studies had to include focus on the workings of TNF-α and investigate the role of this cytokine family in anti-cancer therapy. Articles that did not meet this criterion; articles focusing on other mechanisms of action, were discarded.

### Exclusion criteria

- Correspondences, editorials, commentaries, and pictorial essays
- Case-studies/case-series
- Tertiary and grey literature
- Not published in a Peer-Reviewed Journal
- Not published in the English language

### Inclusion criteria

- Primary articles
- Original research
- Literature-, narrative-, systematic- or scoping review
- Meta-analysis
- Published in Peer-Reviewed Journal
- Published in English.

Review articles were included to get a better understanding of what has already been summarized up until this point, and to provide a background and context for the primary research articles. Original research alone would not be enough to get a complete picture of TNF-α and bacteriotherapy in cancer, even for this small summary. The review articles provide context and the original research articles fill in with details and examples. This gives a broader understanding.

## Results

**Figure 1.**
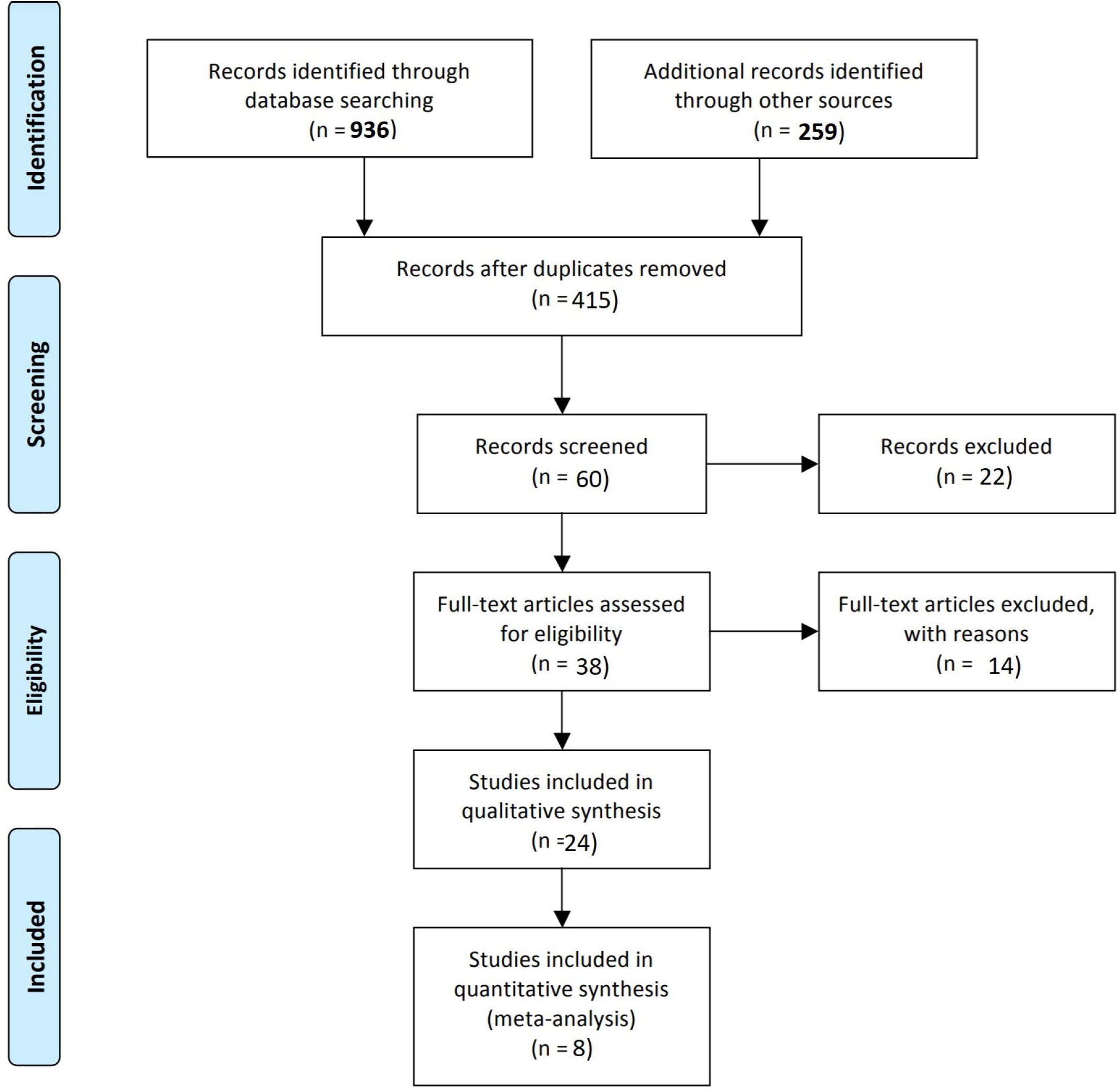
Flow diagram.

Out of 24 interesting and relevant articles 8 were original research articles and 16 provided context and background for the study. Different strains of *Clostridia, Lactococcus, Bifidobacteria, Shigella, Vibrio, Listeria, Escherichia*, and *Salmonella* have been evaluated for cancer therapy as they have different anti-tumour properties. *Bifidobacteria* and *Clostridium* are capable of colonizing hypoxic areas of the tumour, but attenuated and mutated *Salmonella* seem to be the most effective for most types of cancer cells. Another desirable quality is the potential for biological engineering, because if bacteria can be designed for a specific type of cancer, they can be both more effective as well as less harmful for the patient.

**Table 1.**
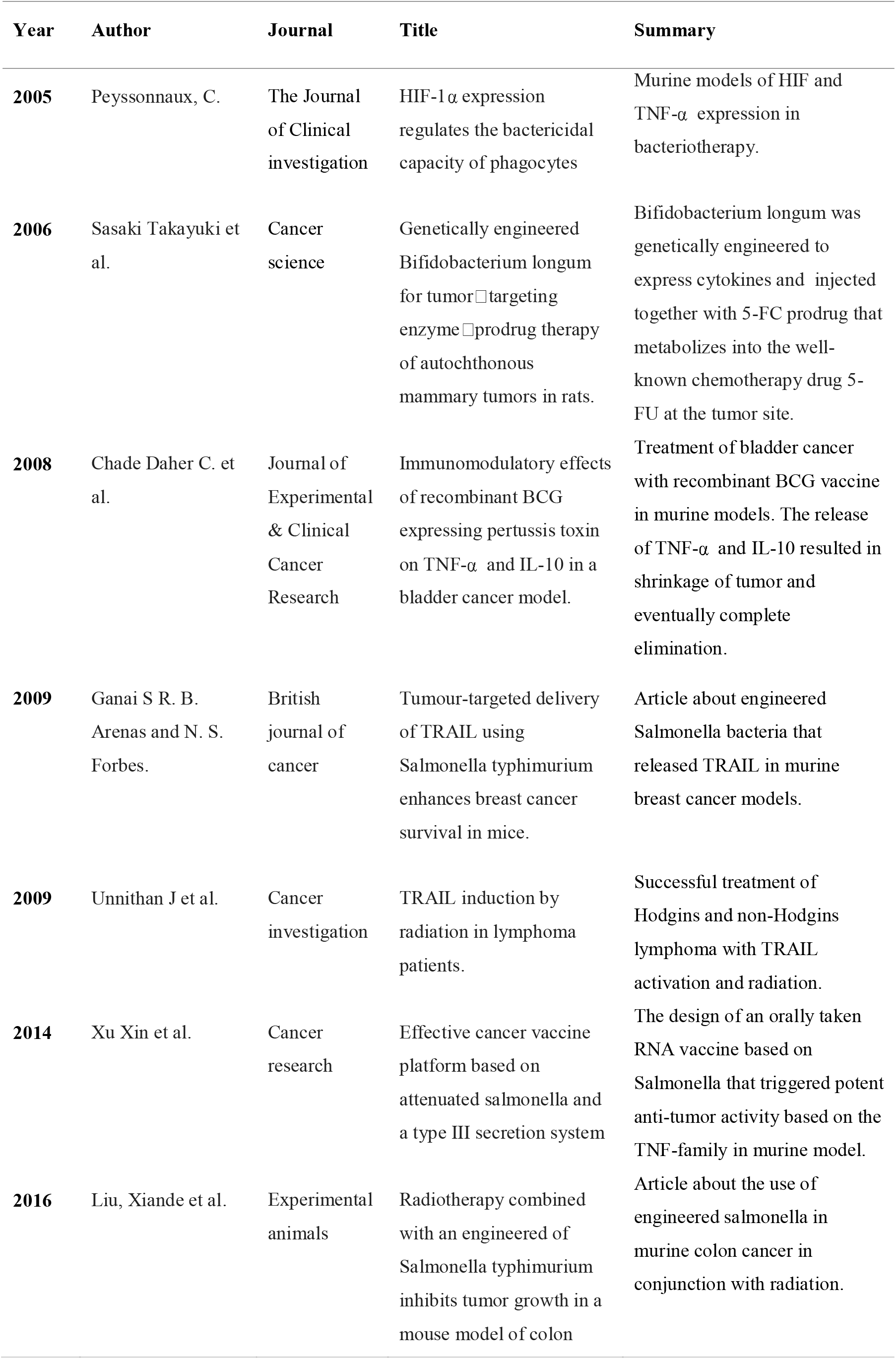
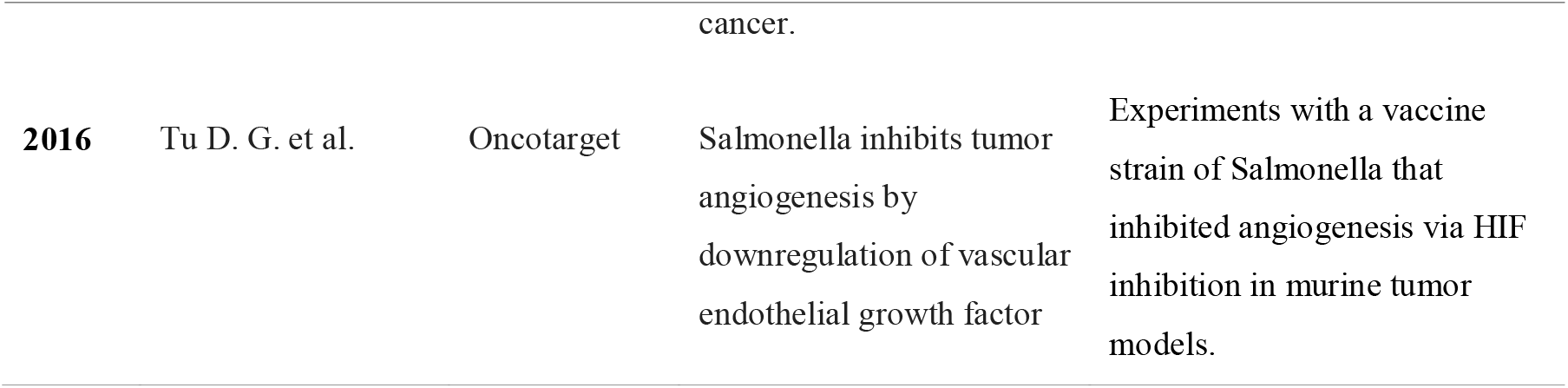
Overview and summary of the original research articles.

In the study with TRAIL-induction via irradiation in patients with Hodgkin’s and non-Hodgkin’s lymphoma good results were seen with significant tumor shrinkage.[22] The idea is that this effect gets enhanced if bacteria contribute to more TRAIL by releasing it to the tumor site. In the study with murine breast cancer models TRAIL-bearing *Salmonella* combined with radiation significantly reduced the risk of death by 76% compared to radiation only. Repeated dosing with TRAIL-bearing *Salmonella* in conjunction with radiation improved the 30-day survival from 0 to 100%. [23]

The immune system of the patient is heavily affected during exposure to bacteria which activates TNF-α as well as other pro-inflammatory cytokines. TNF-α is then drawn to the tumor site because anaerobic bacteria locate there as they seek out a hypoxic area to multiply. Hypoxia is a characteristic feature of the tissue microenvironment during both bacterial infection and tumor development. [18] When bacteria are engineered to trigger local TNF-α release and the effect is enhanced by radiation, TRAIL activates in the bacteria inducing apoptosis in its surroundings.[22]

The study with multiple murine models of melanoma, bladder-, pancreatic-, colon cancer and lymphoma was investigating the possibilities of creating a cancer vaccine by developing a long-lasting protective memory response. The approach was curative in one lymphoma model and all cured animals showed increased cytokine response and formation of memory CD8 T-cells.[6]

The recombinant BCG treatment of bladder cancer was another example of vaccine like treatment and has proven to be superior to chemotherapy. Increase in TNF-α and interleukins (mostly Il-10) leads to memory cell formation and eventually cure if the cancer has not spread to surrounding tissues and organs. [11]

*Bifidobacterium longum* is non-pathogenic and anaerobic which make up an interesting bacteriotherapy candidate. It is potentially safer than *Salmonella* but needs to be genetically engineered to express cytokines and subsequently be paired with the non-toxic pro-drug 5-FC (5-fluorocytosine) that is metabolized into the well know chemotherapy drug 5-FU (5-fluororacil). This combination has been observed to be effective and safe compared to the highly toxic, standard 5-FU treatment.[16]

*Salmonella* has proven its effectiveness in bacteriotherapy due to its aggressive proliferation in anaerobic environments. This inhibits the expression of HIF, thus halts angiogenesis.Targeting angiogenesis was one of the more successful mechanism of tumor inhibition alongside utilization of the TNF superfamily.[18] Several combinations with TNF-α bearing bacteria has been investigated and both TNF-α and TRAIL seem to be effective especially in combination with radiation. [22,23,24]

## Limitations

The scope of this review was very narrow and search results limited because research on oncolytic bacterial therapy with TNF-α as mechanism of action, is not common. Both bacteriotherapy and TNF-α has been studied in multiple contexts including cancer treatment, but seldom in conjunction with each other.

Even though a thorough online search was performed, it is highly likely that retrieval of research papers is incomplete.

## Discussion

The exact mechanism of how bacteria find and invade tumors is still unclear, and injecting bacteria intravenously (in the same manner as Coley’s toxin) is unsafe unless the bacteria has undergone the process of attenuation or some more advanced genetic engineering. A sudden influx of bacteria in the blood stream activates the patient’s immune system and the anaerobic nature of bacteria attracts them to hypoxic tumor sites where TNF-α as well as other pro-inflammatory cytokines follow suit. The systemic inflammatory response creates an influx of blood into the tumor due to vascular disruption, which is followed by necrosis formation, increased bacterial growth, and infiltration of neutrophilic granulocytes (a type of white blood cell). Another mechanism of action is when bacteria are designed to express TNF-α locally in the tumor instead of TNF-α coming from the innate immune system.

Experiments with a genetically engineered type of TRAIL-bearing salmonella has been successfully performed with and without radiation, and it seem to be successful because it mimics the natural behaviour of tumors when they over-express proteins that induce cell death. Apoptosis of tumor cells is desirable in general but it can unfortunately lead to resistance to chemotherapy because the tumor cuts itself off from all blood supply. This is where bacteriotherapy can be extra useful as bacteria can reach these hypoxic regions where pharmaceuticals are unable to reach. TRAIL is more toxic to cancer cells than normal cells but is preferably delivered via genetically engineered bacteria rather than from a systemic immune reaction.

Irradiation is effective because it specifically causes a delay in tumour growth while TRAIL triggers multiple apoptosis mechanism. This results in two-step system of slowing down and then killing the cancer cells.

The relevant original articles focused on *Salmonella, Bifidobacterium*, and *Mycobacterium bovis*. Even though *E. coli* and *Clostridium* also show promising results in bacteriotherapy they have not been studied with TNF-α as main mechanism of action. Best results seem to occur when members of the TNF superfamily locally attack the tumor aided by localized radiation and biologically engineered strains of *Salmonella*. This method seems to yield the best results in a variety of animal models ranging from melanoma to breast cancer, colon cancer and bladder cancer. Not all bacteriotherapy are administered intravenously, some are even taken orally which triggers the immune system differently compared to injectables.

*Salmonella* is so far the most beneficial candidate for bacteriotherapy because of its tendency to proliferate aggressively in anaerobic environments thus inhibits the expression of HIF which halts angiogenesis crucial in tumor progression.

Although these therapies have great potential in tailored cancer treatments, their clinical application is not yet in use, except for one rare exception (BCG for bladder cancer). Biological therapies continue to show promising results in animal studies, but the main bottleneck for clinical application is to determine the best tumor colonizing bacterial host as well as assessing the therapeutic efficacy either as single treatment or in combination with other therapies. Bacterial vectors are more difficult to manage than pharmaceutical therapies thus limited to early studies. Pharmaceuticals are generally more exact, easily administrated and the dose can be calculated exactly. Bacteriotherapy is generally more like a vaccine, which is desirable, but it is hard to predict how each individual immune system will react to the infection. The risk of cytokine release syndrome is a serious fatal side-effect of systemic inflammation. Development of genetically engineered strains might counteract this risk, making bacteriotherapy a safer alternative than chemotherapy in the future. However

## Conclusions

This review gives a short overview of the current status of TNF-α as the main mechanism of action in oncogenic bacteriotherapy. Although there have been many promising results, most studies on bacteriotherapy for the treatment of cancer have been stopped in the in vitro stages and only a few studies have gone from in vitro to clinical trial, registration and use as medicines. Small phase 1 studies have been performed but this field of anti-cancer treatments needs further in vivo research, as well as clinical trials. The multifactorial physiology of cancer makes the treatment difficult as it varies greatly depending on the volume, stage and site of the tumour as well as if metastasis has occurred or not. Chemotherapy, radiotherapy and immunotherapy can all be subject to resistance thus be ineffective and lead to more side effects than benefits. Especially in the later stages of the treatment which is the biggest reason for the development of complimentary therapies.

## Data Availability

N/A

## Declarations

### Ethics approval and consent to participate

N/A

### Consent for publication

N/A

### Availability of data and material

N/A

## Author Contributions

Martina Johansson came up with the idea of the review, performed the search and analysed the retrieved papers. Fredrik H Nystrom reviewed and supervised the writing process.

## Acknowledgements

N/A

## References

1. Zidi, Inès, et al. “TNF-α and its inhibitors in cancer.” Medical Oncology 27.2 (2010): 185–198.

2. Cai, Weibo, et al. “Targeted cancer therapy with tumor necrosis factor-α .” Biochemistry insights 1 (2008): BCI–S901.

3. Nallar, Shreeram C., De-Qi Xu, and Dhan V. Kalvakolanu. “Bacteria and genetically modified bacteria as cancer therapeutics: Current advances and challenges.” Cytokine 89 (2017): 160–172.

4. Mayes, Patrick A., Kenneth W. Hance, and Axel Hoos. “The promise and challenges of immune agonist antibody development in cancer.” Nature Reviews Drug Discovery 17.7 (2018): 509–527.

5. Wei, Ming Q., et al. “Facultative or obligate anaerobic bacteria have the potential for multimodality therapy of solid tumours.” European journal of cancer 43.3 (2007): 490–496.

6. Xu, Xin, et al. “Effective cancer vaccine platform based on attenuated salmonella and a type III secretion system.” Cancer research 74.21 (2014): 6260–6270.

7. McCarthy, E. F. (2006). The toxins of William B. Coley and the treatment of bone and soft-tissue sarcomas. The Iowa orthopaedic journal, 26, 154.

8. Richardson, Mary Ann, et al. “Coley toxins immunotherapy: a retrospective review.” Alternative therapies in health and medicine 5.3 (1999): 42.

9. Carlson, Robert D., John C. Flickinger, and Adam E. Snook. “Talkin’Toxins: From Coley’s to Modern Cancer Immunotherapy.” Toxins 12.4 (2020): 241.

10. Najafi, M., Hashemi Goradel, N., Farhood, B., Salehi, E., Nashtaei, M. S., Khanlarkhani, N., … & Kashani, I. R. (2019). Macrophage polarity in cancer: A review. Journal of cellular biochemistry, 120(3), 2756–2765.

11. Chade, Daher C., et al. “Immunomodulatory effects of recombinant BCG expressing pertussis toxin on TNF-α and IL-10 in a bladder cancer model.” Journal of Experimental & Clinical Cancer Research 27.1 (2008): 78.

12. Taniguchi, Shun’ichiro, et al. “Targeting solid tumours with nonJpathogenic obligate anaerobic bacteria.” Cancer science 101.9 (2010): 1925–1932.

13. St Jean, A. T., Zhang, M., & Forbes, N. S. (2008). Bacterial therapies: completing the cancer treatment toolbox. Current opinion in biotechnology, 19(5), 511–517.

14. Patyar, S., Joshi, R., Byrav, D. P., Prakash, A., Medhi, B., & Das, B. K. (2010). Bacteria in cancer therapy: a novel experimental strategy. Journal of biomedical science, 17(1), 21.

15. Lemmon, M. J., Van Zijl, P., Fox, M. E., Mauchline, M. L., Giaccia, A. J., Minton, N. P., & Brown, J. M. (1997). Anaerobic bacteria as a gene delivery system that is controlled by the tumor microenvironment. Gene therapy, 4(8), 791–796.

16. Sasaki, T., Fujimori, M., Hamaji, Y., Hama, Y., Ito, K. I., Amano, J., & Taniguchi, S.(2006). Genetically engineered Bifidobacterium longum for tumorJtargeting enzymeJprodrug therapy of autochthonous mammary tumors in rats. Cancer science, 97(7), 649–657.

17. Tu, D. G., Chang, W. W., Lin, S. T., Kuo, C. Y., Tsao, Y. T., & Lee, C. H. (2016). Salmonella inhibits tumor angiogenesis by downregulation of vascular endothelial growth factor. Oncotarget, 7(25), 37513.

18. Peyssonnaux, C., Datta, V., Cramer, T., Doedens, A., Theodorakis, E. A., Gallo, R. L.,… & Johnson, R. S. (2005). HIF-1α expression regulates the bactericidal capacity of phagocytes. The Journal of clinical investigation, 115(7), 1806–1815.

19. Wajant, H., K. Pfizenmaier, and P. Scheurich. “TNF-α related apoptosis inducing ligand (TRAIL) and its receptors in tumour surveillance and cancer therapy.” Apoptosis 7.5 (2002): 449–459.

20. Palazon, A., Goldrath, A. W., Nizet, V., & Johnson, R. S. (2014). HIF transcription factors, inflammation, and immunity. Immunity, 41(4), 518–528.

21. Khare, Ritu, Robert Leaman, and Zhiyong Lu. “Accessing biomedical literature in the current information landscape.” Biomedical Literature Mining. Humana Press, New York, NY, 2014. 11–31.

22. Liu, Xiande, et al. “Radiotherapy combined with an engineered of Salmonella typhimurium inhibits tumour growth in a mouse model of colon cancer.” Experimental animals (2016): 16–0033

23. Unnithan, J., & Macklis, R. M. (2004). TRAIL induction by radiation in lymphoma patients. Cancer investigation, 22(4), 522–525.

24. Ganai, S., R. B. Arenas, and N. S. Forbes. “Tumour-targeted delivery of TRAIL using Salmonella typhimurium enhances breast cancer survival in mice.” British journal of cancer 101.10 (2009): 1683–1691

